# Updated estimates of comorbidities associated with risk for COVID-19 complications based on US data

**DOI:** 10.1101/2020.05.02.20088781

**Authors:** Mary L. Adams, Joseph Grandpre, David L. Katz

## Abstract

We updated previous estimates (wwwnc.cdc.gov/eid/article/26/8/20-0679_article) of adults with any underlying condition increasing risk of complications from COVID-19 using recent US hospitalization data instead of mortality data from China. This substitutes obesity for cancer in the definition and increased the percentage of adults reporting ≥1 condition to 56.0% (95% CI 55.7-56.4). When controlled for all measures listed, factors increasing odds of reporting any of the underlying conditions include being male, older, African American, American Indian, household income <$25,000, < high school education, underinsurance, living in the South or Midwest (vs. West), plus the risk factors of ever smoking, sedentary lifestyle, and inadequate fruit and vegetable consumption. Population-attributable risk for the listed risk factors was 13.0%, 12.6%, and 15.0% respectively. Results have potential implications for policies based on risk-stratification of the population and for improvement of risk status through lifestyle change. National support for a “health promotion” campaign would be timely.

---

One of the novel aspects of the current coronavirus pandemic is that information is constantly being updated as more studies are published. We had earlier reported estimates of comorbidities affecting risk for complications from COVID-19 that were based on mortality data from China (*1*). Newly released US data on hospitalizations for COVID-19 from 14 states indicate a slightly different list of conditions affecting risk, with obesity added and cancer subtracted (*2*). Those preliminary results indicate that 89% of those hospitalized for COVID-19 had an underlying condition, the most common being hypertension, obesity, chronic lung disease (including asthma and chronic obstructive pulmonary disease), diabetes, and cardiovascular disease (*2*). Similar results were found for ICU admissions on a smaller sample and without obesity (*3*). The Guidelines for Opening up America Again released by the White House on April 16, 2020 (*4*) include a similar list to define vulnerable individuals, except that list includes elderly (undefined) and those with a compromised immune system and omits cardiovascular disease.

Our main objective for this study was to use population based US data to estimate the fraction of adults in the community who might be a risk of hospitalization for COVID-19 (2) due to reporting any of the most common underlying conditions listed above. Secondly, we sought to determine the demographic and risk factors that increased the likelihood of having one or more of these underlying conditions. And thirdly, using the information about potentially modifiable risk factors obtained through our second objective, we estimated population attributable-risk for conditions increasing risk for COVID-19 hospitalization.

## Methods

We used publicly available 2017 Behavioral Risk Factor Surveillance System (BRFSS) data (*5*) from telephone surveys of 444,649 randomly selected adults ages 18 and older in the 50 states and the District of Columbia (DC). The BRFSS includes only non-institutionalized adults so residents of prisons, nursing homes and assisted living facilities are among those not surveyed. We chose to use 2017 data in order to include hypertension which was not addressed in 2018. Data were adjusted for the probability of selection and weighted to be representative of the adult population in each state by age, gender, race/ethnicity, marital status, education, home ownership, and telephone type and included weights and stratum variables needed for analysis. Results were not age adjusted in order to reflect the age distribution of each state rather than a standard population. The median response rate for cell phone and land line surveys combined was 47.2%, ranging from 33.9% in California to 61.1% in Utah (*6*). Reliability and validity of the BRFSS have been found to be moderate to high for many survey measures, in particular those used here which can be checked versus medical records (*7*).

The key variable was a composite measure including adults reporting they were ever told they had diabetes (excluding if only when pregnant), asthma (and still had it), chronic obstructive pulmonary disease (COPD), hypertension, cardiovascular disease (CVD; ever told they had a heart attack, angina, coronary heart disease, or a stroke) or they were obese with body mass index ≥30.0 based on self-reported height and weight. The number of conditions was counted for each respondent and adults who reported at least one were considered to be at risk of hospitalization due to an underlying condition. Risk factors included ever smoking 100 cigarettes, sedentary lifestyle, defined as no leisure time physical activity in the past month, and inadequate fruit and vegetable consumption, defined as consuming the combination <5 times per day based on responses to five separate questions, with responses to the question about fried potatoes or French fries excluded. A composite measure was also created for risk factors by counting the number for each respondent and determining those with each number of risks from 0-3.

Demographic measures included age group (18-29, 30-39, 40-49, 50-59, 60-69, 70-79, and 80+ years, which was created by combining 5 year age groups provided in the data set), self-reported race/ethnicity (non-Hispanic white, Black or African American, Hispanic of any race, American Indian/Alaska native, Asian/Pacific Islander, and other), less than a high school education (vs. high school graduate or higher), household income < $25,000 (vs. all other incomes including unknown), being underinsured (having no health insurance coverage or reporting a time in the past year when the needed health care but could not get it due to cost), state of residence which included DC, whether or not they lived within the center city of a Metropolitan Statistical Area (MSA), and census region (Northeast, Midwest, South, or West)(*8*).

Stata version 14.1 (Stata Corp LP, College Station, TX) was used for analysis to account for the complex sample design of the BRFSS. Point estimates and 95% confidence intervals are reported using the weights, stratum, and PSU variables supplied in the data set (*5*). Missing values were excluded from analysis. Population-attributable risk (PAR) was used to compare the contributions of individual risk factors to the composite measure of any of the 6 underlying conditions and to each of the 6 component conditions (*9*). PAR estimates take into account not only the relative risk of people with that risk factor developing the outcome, but also the prevalence of the risk factor in the population (*10*). PARs were estimated in Excel using Levin’s formula (*11*) and adjusted odds ratios (AOR) obtained from logistic regression instead of unadjusted relative risk.

## Results

Sample was described earlier (*1*). Overall, 56.0% (95% CI 55.7-56.4) of respondents had one or more of the underlying conditions, ranging from 33.7% among 18-29 year olds to 79.1% for those 80 years and older. Prevalence rates of the separate conditions were 8.5% for CVD, 6.6% for COPD, 9.1% for asthma, 10.8% for diabetes, 32.4% for hypertension, and 30.1% for obesity. While the percentage of adults with any of the conditions increased with age (Table 1), 60.7% of the total was younger than age 60. Rates also varied by all the measures in Table 1 except underinsurance. State rates ranged from 45.6% in DC to 68.8% in West Virginia. Using the definition for vulnerable individuals according to the Guidelines for Opening up America (*4*) which adds age (we used age 65 as elderly) and removes CVD, that prevalence rate is 60.4% (95% CI 60.0-60.7).

**Table 1.**
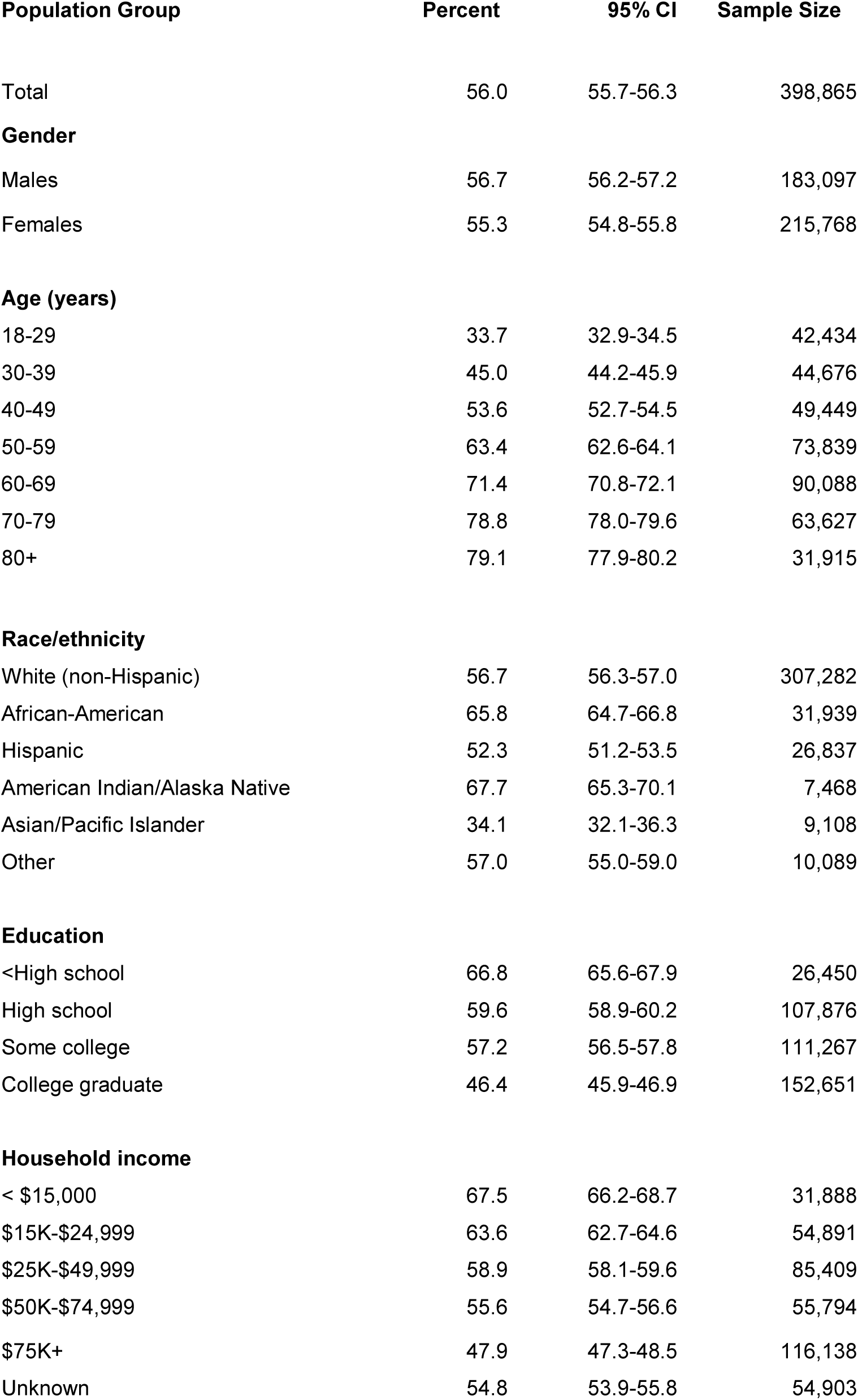
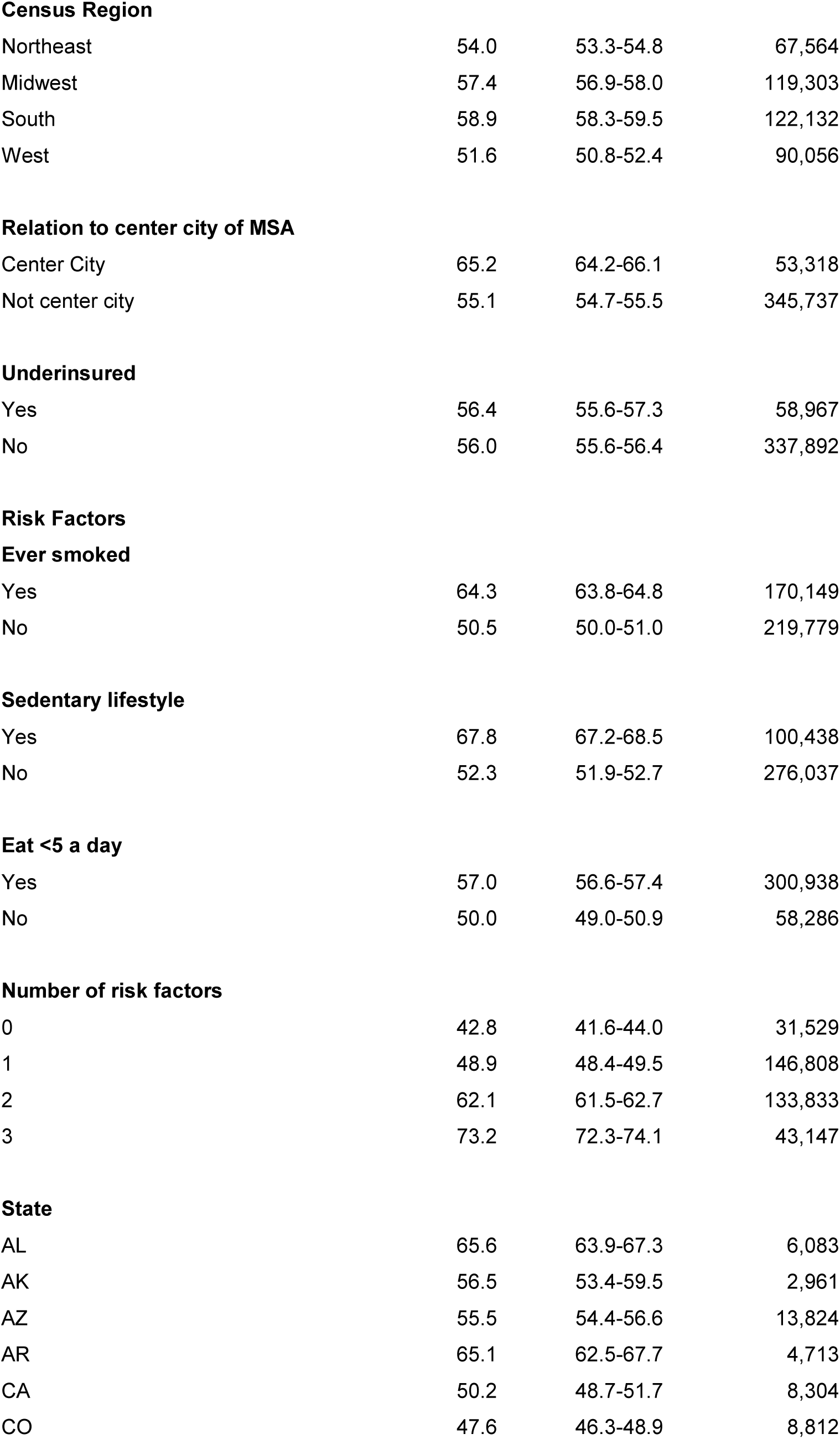
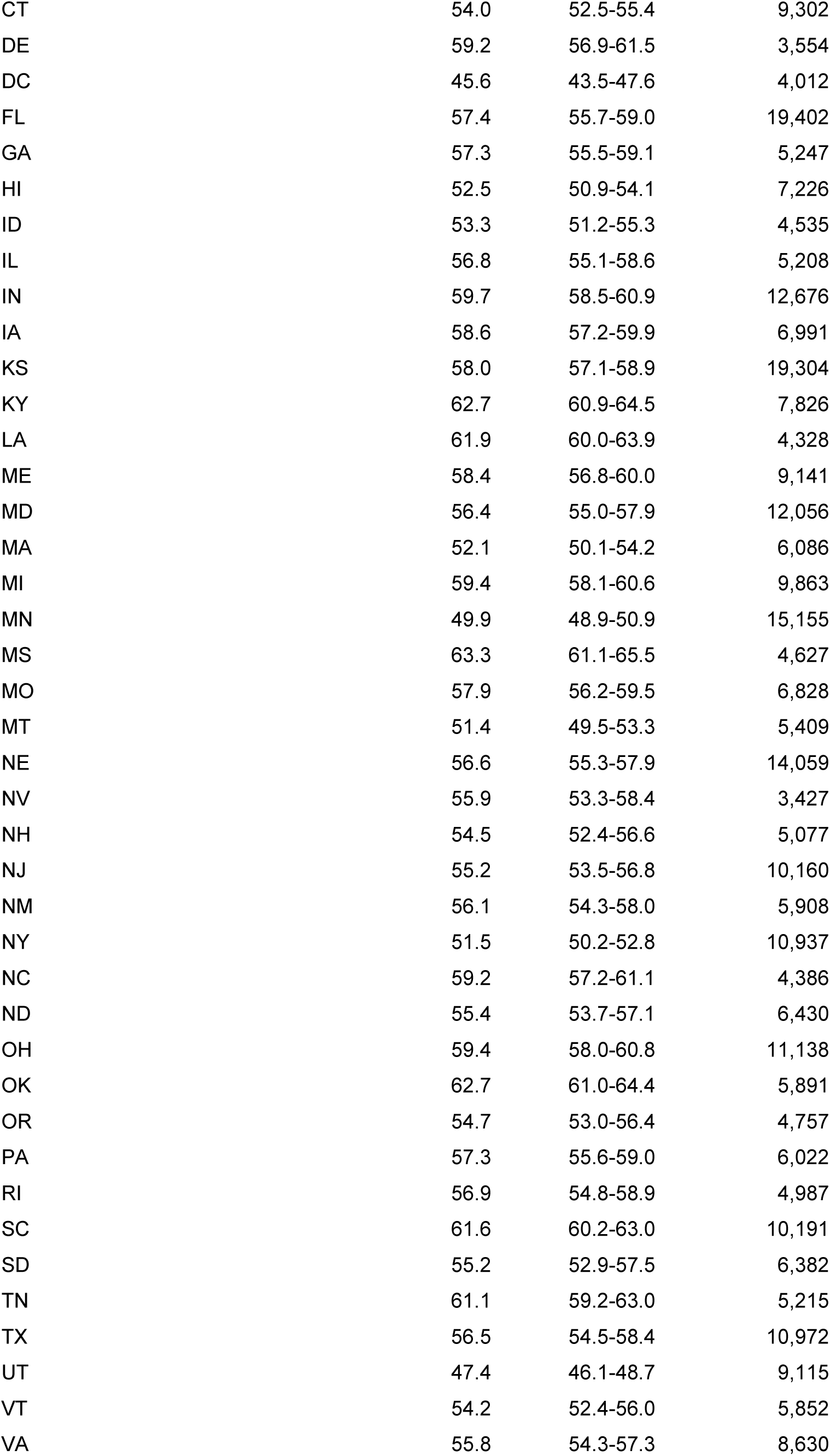
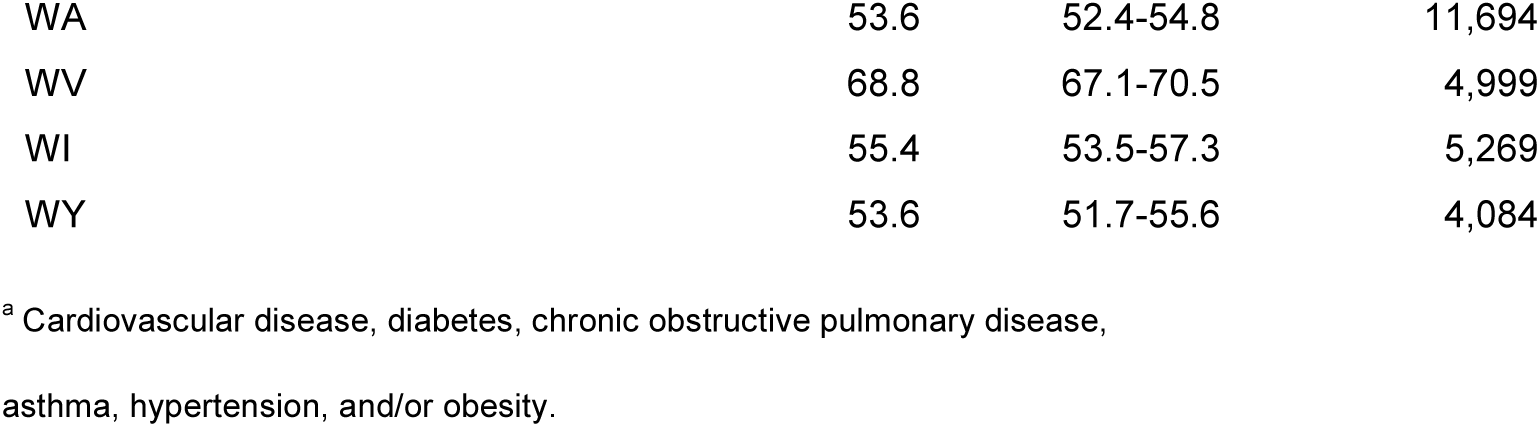
Demographics of adults with any of 6 underlying conditions^a^; 2017 Behavioral Risk Factor Surveillance System.

| **Population Group** | **Percent** | **95% CI** | **Sample Size** |
| --- | --- | --- | --- |
| Total | 56.0 | 55.7-56.3 | 398,865 |
| **Gender** |  |  |  |
| Males | 56.7 | 56.2-57.2 | 183,097 |
| Females | 55.3 | 54.8-55.8 | 215,768 |
| **Age (years)** |  |  |  |
| 18-29 | 33.7 | 32.9-34.5 | 42,434 |
| 30-39 | 45.0 | 44.2-45.9 | 44,676 |
| 40-49 | 53.6 | 52.7-54.5 | 49,449 |
| 50-59 | 63.4 | 62.6-64.1 | 73,839 |
| 60-69 | 71.4 | 70.8-72.1 | 90,088 |
| 70-79 | 78.8 | 78.0-79.6 | 63,627 |
| 80+ | 79.1 | 77.9-80.2 | 31,915 |
| **Race/ethnicity** |  |  |  |
| White (non-Hispanic) | 56.7 | 56.3-57.0 | 307,282 |
| African-American | 65.8 | 64.7-66.8 | 31,939 |
| Hispanic | 52.3 | 51.2-53.5 | 26,837 |
| American Indian/Alaska Native | 67.7 | 65.3-70.1 | 7,468 |
| Asian/Pacific Islander | 34.1 | 32.1-36.3 | 9,108 |
| Other | 57.0 | 55.0-59.0 | 10,089 |
| **Education** |  |  |  |
| <High school | 66.8 | 65.6-67.9 | 26,450 |
| High school | 59.6 | 58.9-60.2 | 107,876 |
| Some college | 57.2 | 56.5-57.8 | 111,267 |
| College graduate | 46.4 | 45.9-46.9 | 152,651 |
| **Household income** |  |  |  |
| < \$15,000 | 67.5 | 66.2-68.7 | 31,888 |
| \$15K-\$24,999 | 63.6 | 62.7-64.6 | 54,891 |
| \$25K-\$49,999 | 58.9 | 58.1-59.6 | 85,409 |
| \$50K-\$74,999 | 55.6 | 54.7-56.6 | 55,794 |
| \$75K+ | 47.9 | 47.3-48.5 | 116,138 |
| Unknown | 54.8 | 53.9-55.8 | 54,903 |
| **Census Region** |  |  |  |
| Northeast | 54.0 | 53.3-54.8 | 67,564 |
| Midwest | 57.4 | 56.9-58.0 | 119,303 |
| South | 58.9 | 58.3-59.5 | 122,132 |
| West | 51.6 | 50.8-52.4 | 90,056 |
| **Relation to center city of MSA** |  |  |  |
| Center City | 65.2 | 64.2-66.1 | 53,318 |
| Not center city | 55.1 | 54.7-55.5 | 345,737 |
| **Underinsured** |  |  |  |
| Yes | 56.4 | 55.6-57.3 | 58,967 |
| No | 56.0 | 55.6-56.4 | 337,892 |
| **Risk Factors****Ever smoked** |  |  |  |
| Yes | 64.3 | 63.8-64.8 | 170,149 |
| No | 50.5 | 50.0-51.0 | 219,779 |
| **Sedentary lifestyle** |  |  |  |
| Yes | 67.8 | 67.2-68.5 | 100,438 |
| No | 52.3 | 51.9-52.7 | 276,037 |
| **Eat <5 a day** |  |  |  |
| Yes | 57.0 | 56.6-57.4 | 300,938 |
| No | 50.0 | 49.0-50.9 | 58,286 |
| **Number of risk factors** |  |  |  |
| 0 | 42.8 | 41.6-44.0 | 31,529 |
| 1 | 48.9 | 48.4-49.5 | 146,808 |
| 2 | 62.1 | 61.5-62.7 | 133,833 |
| 3 | 73.2 | 72.3-74.1 | 43,147 |
| **State** |  |  |  |
| AL | 65.6 | 63.9-67.3 | 6,083 |
| AK | 56.5 | 53.4-59.5 | 2,961 |
| AZ | 55.5 | 54.4-56.6 | 13,824 |
| AR | 65.1 | 62.5-67.7 | 4,713 |
| CA | 50.2 | 48.7-51.7 | 8,304 |
| CO | 47.6 | 46.3-48.9 | 8,812 |
| CT | 54.0 | 52.5-55.4 | 9,302 |
| DE | 59.2 | 56.9-61.5 | 3,554 |
| DC | 45.6 | 43.5-47.6 | 4,012 |
| FL | 57.4 | 55.7-59.0 | 19,402 |
| GA | 57.3 | 55.5-59.1 | 5,247 |
| HI | 52.5 | 50.9-54.1 | 7,226 |
| ID | 53.3 | 51.2-55.3 | 4,535 |
| IL | 56.8 | 55.1-58.6 | 5,208 |
| IN | 59.7 | 58.5-60.9 | 12,676 |
| IA | 58.6 | 57.2-59.9 | 6,991 |
| KS | 58.0 | 57.1-58.9 | 19,304 |
| KY | 62.7 | 60.9-64.5 | 7,826 |
| LA | 61.9 | 60.0-63.9 | 4,328 |
| ME | 58.4 | 56.8-60.0 | 9,141 |
| MD | 56.4 | 55.0-57.9 | 12,056 |
| MA | 52.1 | 50.1-54.2 | 6,086 |
| MI | 59.4 | 58.1-60.6 | 9,863 |
| MN | 49.9 | 48.9-50.9 | 15,155 |
| MS | 63.3 | 61.1-65.5 | 4,627 |
| MO | 57.9 | 56.2-59.5 | 6,828 |
| MT | 51.4 | 49.5-53.3 | 5,409 |
| NE | 56.6 | 55.3-57.9 | 14,059 |
| NV | 55.9 | 53.3-58.4 | 3,427 |
| NH | 54.5 | 52.4-56.6 | 5,077 |
| NJ | 55.2 | 53.5-56.8 | 10,160 |
| NM | 56.1 | 54.3-58.0 | 5,908 |
| NY | 51.5 | 50.2-52.8 | 10,937 |
| NC | 59.2 | 57.2-61.1 | 4,386 |
| ND | 55.4 | 53.7-57.1 | 6,430 |
| OH | 59.4 | 58.0-60.8 | 11,138 |
| OK | 62.7 | 61.0-64.4 | 5,891 |
| OR | 54.7 | 53.0-56.4 | 4,757 |
| PA | 57.3 | 55.6-59.0 | 6,022 |
| RI | 56.9 | 54.8-58.9 | 4,987 |
| SC | 61.6 | 60.2-63.0 | 10,191 |
| SD | 55.2 | 52.9-57.5 | 6,382 |
| TN | 61.1 | 59.2-63.0 | 5,215 |
| TX | 56.5 | 54.5-58.4 | 10,972 |
| UT | 47.4 | 46.1-48.7 | 9,115 |
| VT | 54.2 | 52.4-56.0 | 5,852 |
| VA | 55.8 | 54.3-57.3 | 8,630 |
| WA | 53.6 | 52.4-54.8 | 11,694 |
| WV | 68.8 | 67.1-70.5 | 4,999 |
| WI | 55.4 | 53.5-57.3 | 5,269 |
| WY | 53.6 | 51.7-55.6 | 4,084 |

<sup>a</sup> Cardiovascular disease, diabetes, chronic obstructive pulmonary disease,
asthma, hypertension, and/or obesity.

Prevalence rates for the risk factors used in PAR estimates for all ages were 40.4% for ever smoking, 26.6% for sedentary lifestyle, and 84.1% for inadequate fruit and vegetable consumption. Among ever smokers of all ages, 40.7% currently smoke, including 50.6% of those ages 18-59 and 22.8% of adults ages 60 and older. Those estimates result in current smoking rates of 16.5%, 18.6%, and 11.2% respectively.

Results of multiple logistic regression controlled for age, gender, education, income, race/ethnicity, underinsurance, region, and the 3 behavioral risk factors (Table 2) indicate that male gender, increasing age, less education and income, being underinsured, having each of the risk factors, being African American, or American Indian/Alaska Native and living in either the Midwest or South significantly increased the odds of having an underlying condition shown to be associated with COVID-19 hospitalizations. Hispanic ethnicity had no effect while being Asian/Pacific Islander decreased the odds of having any of the conditions. Adding a variable for living in the center city of an MSA had no effect when controlled for the other measures. Including the composite risk factor measure in the logistic regression model in place of the separate risks confirmed the step-wise increase with an AOR of 2.56 (2.38-2.76) for adults with all 3 risk factors compared with those reporting none.

**Table 2.** Multiple logistic regression results for outcome of reporting any of 6 underlying conditions ^a^; 2017 Behavioral Risk Factor Surveillance System; N=346,198.

| **Group** | **Adjusted Odds Ratio** | **95% CI** |
| --- | --- | --- |
| Females (referent) |  |  |
| Males | 1.15 | 1.12-1.19 |
| Age 18-29 (referent) |  |  |
| 30-39 | 1.57 | 1.49-1.67 |
| 40-49 | 2.21 | 2.09-2.34 |
| 50-59 | 3.27 | 3.10-3.46 |
| 60-69 | 4.79 | 4.53-5.06 |
| 70-79 | 7.19 | 6.73-7.68 |
| 80+ | 7.01 | 6.34-7.76 |
| Never smoked (referent) |  |  |
| Ever smoked | 1.37 | 1.33-1.42 |
| Not sedentary (referent) |  |  |
| Sedentary | 1.54 | 1.48-1.60 |
| Fruit & vegetables 5 X/day (referent) |  |  |
| < 5 time/day | 1.21 | 1.16-1.26 |
| High school grad/GED (referent) |  |  |
| Not high school graduate | 1.23 | 1.14-1.31 |
| Income \$25,000+ (referent) | | |
| Income <\$25,000 | 1.46 | 1.40-1.53 |
| Not underinsured (referent) |  |  |
| Underinsured | 1.11 | 1.06-1.16 |
| White non-Hispanic (referent) |  |  |
| African American | 1.67 | 1.57-1.76 |
| Hispanic any race | 1.03 | 0.97-1.10 |
| American Indian/AK Native | 1.67 | 1.46-1.92 |
| Asian/Pacific Islander | 0.57 | 0.51-0.63 |
| Other | 1.27 | 1.15-1.40 |
| West region (referent) |  |  |
| Northeast | 0.98 | 0.94-1.03 |
| Midwest | 1.14 | 1.09-1.19 |
| South | 1.13 | 1.08-1.19 |

<sup>a</sup> Cardiovascular disease, diabetes, chronic obstructive pulmonary disease,
asthma, hypertension, and/or obesity.

PAR estimates for the 3 potentially modifiable risk factors, estimated using the AORs in Table 2 are shown in Table 3. Among all adults, these 3 risk factors were estimated to account for 40.6% of the total risk of any underlying condition, 34.2% of total risk for adults 18-59 and 59.7% of total risk for adults age 60 and older. The total attributable risk for all adults for the separate conditions ranged from 18.7% for asthma to 69.2% for COPD. (Table 3).

**Table 3.** Population attributable-risk (PAR) for selected outcomes. 2017 Behavioral Risk Factor Surveillance System; Maximum N=372,041

| **Outcome** | **PAR**<br>Ever<br>smoked | **PAR**<br>Sedentary<br>lifestyle | **PAR**<br>Fruits/vegetables<br>< 5X/day | **Sum**<br>**of**<br>**PARs** |
| --- | --- | --- | --- | --- |
| Any of 6 conditions<sup>a</sup> all adults | 13.0% | 12.6% | 15.0% | 40.6% |
| Any of 6 conditions<sup>a</sup> age <60 | 12.6% | 9.8% | 11.8% | 34.2% |
| Any of 6 conditions<sup>a</sup> age 60+ | 13.6% | 21.6% | 24.4% | 59.7% |
| Diabetes | 5.0% | 11.5% | 15.6% | 32.1% |
| Hypertension | 8.5% | 8.5% | 13.1% | 30.2% |
| COPD | 52.5% | 16.6% | 0.0% | 69.2% |
| Asthma | 11.8% | 6.9% | 0.0% | 18.7% |
| CVD | 22.3% | 8.7% | 0.0% | 31.0% |
| Obesity | 0.0% | 14.5% | 13.8% | 28.3% |

Abbreviations: PAR: population attributable risk; COPD: chronic obstructive pulmonary disease; CVD: cardiovascular disease.
<sup>a</sup> Cardiovascular disease, diabetes, chronic obstructive pulmonary disease, asthma, hypertension, and/or obesity, conditions
found to be associated with US hospitalizations for COVID-19.

## Discussion

We estimate that 56.0% of US adults, with a wide range across age groups and states, have one or more underlying conditions that increase risk of hospitalization due to COVID-19. Our previous definition of adults at risk of complications from COVID-19 (*1*) that excluded obesity and added cancer based on data for China where obesity is much less prevalent (*13*) resulted in an estimate of 45.4%. Using the definition of vulnerable individuals in the White House Guidelines (*4*) resulted in an estimate for those at risk of 60.4%. Thus, except for measures that exclude obesity, it appears that over 50% of all adults are at risk for hospitalization or can be considered vulnerable individuals. In addition, while rates increase with increasing age group and age is the key predictor of reporting an underlying condition, over half of adults with underlying conditions using *any* of these definitions are < 60 years. Although our definition of underlying conditions does not include every condition reported among the 89% of persons hospitalized for COVID-19 (*2*) who reported any, our population based prevalence of 56% for any of the conditions is consistent with any of these 6 conditions increasing risk for hospitalization. The underlying chronic conditions increasing risk for COVID-19 hospitalizations are also very similar to those increasing risk for seasonal influenza complications (*14*).

All the underlying conditions used in our measures are conditions for which behavioral risk factors have been well established (*9, 12*). Our list of underlying conditions includes obesity, diabetes, and hypertension, which are also considered risk factors in some studies (*9, 12*). Causal relationships between risk factors and outcomes can be complicated; for example, previous results (*9*) have shown that obesity contributes 37.8% to attributable-risk for diabetes, 30.9% to hypertension and 16.4% to asthma, while hypertension and diabetes contribute 29.3% and 6.7% respectively to CVD. For simplification, we limited our list of risk factors to lifetime smoking, sedentary lifestyle, and inadequate fruit and vegetable consumption. Each selected risk factor was associated with increased likelihood of reporting any of the 6 underlying conditions with AORs of 1.21 to 1.54. In addition, the unadjusted results for increasing number of risk factors indicate a step-wise increase in the percentage of adults reporting any of the underlying conditions with each additional risk factor as shown in Table 1. This result was confirmed by logistic regression and provides new and informative data on factors that predict increased likelihood of reporting any of the underlying conditions.

The results on the risk factors suggest the impact is consequential, with total PARs for the 3 risk factors of 40.6% overall, 34.2% for adults 18-59 and 59.7% for adults ages 60 and older. Of course the smoking measure included lifetime smoking in order to capture effects among former smokers, while current smoking rates are much lower at 16.5% overall. The results suggest the potential for considerable improvement by reducing just these 3 risk factors, or even 2 of the 3. For example, increasing fruit and vegetable consumption and initiating an exercise program such as walking could potentially lower the future risk of developing any of these underlying conditions, with total elimination lowering risk by as much as 46.0% for adults ages 60 and older and 21.6% for those 18-59 years.

In addition to the risk factors mentioned above, a long list of variables increased the likelihood of reporting an underlying condition when controlled for other factors. Foremost among them are being older, male, and African American, all groups which preliminary hospitalization data (*2*) suggested were disproportionately affected by COVID-19. Results from this study indicate that even when controlled for age, sex, underinsurance, income, region, and the 3 risk factors, men, African-Americans, and adults age 60 and older are all more likely to report any of the underlying conditions increasing risk for hospitalizations, with AORs of 1.15, 1.67 and ≥4.79 respectively. Thus it appears part of the reason these groups were disproportionately hospitalized for COVID-19 (*2*) is because they are more likely to report any of the underlying conditions. More study will be needed to determine if other factors increasing likelihood of underlying conditions such as less education and income, and being underinsured or American Indian, are confirmed as increasing hospitalization rates among adults with COVID-19.

In this study we grouped states into regions based on the 4 census regions (*8*) which clearly showed that living in either the Midwest or South increased risk of reporting an underlying condition compared with the West (or Northeast). This result is consistent with studies showing that obesity rates are also highest in these regions (*15*). And while living in a city (center city of an MSA) was associated with reporting an underlying condition in unadjusted analysis, once controlled for demographics and risk factors, urban residence was no longer statistically significant. On the other hand, underinsurance was not associated with reporting an underlying condition in unadjusted analysis but was a statistically significant predictor of reporting an underlying condition once controlled for all measures in the model. Underinsurance has the potential to present a barrier to hospitalization for any reason.

## Limitations

Our study does not address possible differences in contracting the disease, only the risk of hospitalization among those with COVID-19, based on preliminary US results for underlying conditions (*2,3*). Only non-institutionalized adults are surveyed so 1.3 million adults in nursing homes (*16*) were excluded which almost certainly underestimates those with underlying conditions who are included in hospitalization data. Data are self-reported and reliability and validity can vary for different measures tested (*7*). But as long as a respondent was told they had a chronic condition, validity was high. The variable for living in the center city of an MSA was missing for a large number of respondents. Reverse causality may be a factor in some of the PAR estimates, especially for sedentary lifestyle. Age groups used for analysis did not match those used for weighting data but that should have a minimal effect on results. Low response rates could introduce bias but, as noted, validity appears high for the measures used in this study. A dearth of denominator data addressing the actual prevalence of coronavirus infection and immunity in the United States makes it difficult to draw reliable conclusions about overall rates of hospitalization or death among those exposed.

## Conclusion

We estimate 56.0% of US adults are at risk of needing hospitalization for COVID-19 due to underlying conditions. Such estimates will vary depending on exact criteria used to define underlying conditions but represent a substantial fraction of all US adults. These underlying conditions are, in turn, associated with modifiable risk factors including ever smoking, being sedentary and inadequate fruit and vegetable consumption. The three risk factors were estimated to contribute 40.6% of attributable-risk for all adults reporting any of the underlying conditions in this study including 34.2% of attributable-risk among adults ages 18-59 and 59.7% for adults ages 60 and older. These results suggest the potential for policies based on risk-stratification of the population and for possible improvement of risk status through lifestyle change. A national focus on, and support for, a “health promotion” campaign would be timely.

## Data Availability

Survey data are available at:
https://www.cdc.gov/brfss/annual_data/annual_2017.html

## Funding

Data collection, analysis, and interpretation of data for this study were supported by the Centers for Disease Control and Prevention (CDC) Grant/Cooperative Agreement number 1U58DP006069-01.

## Notes

### Competing Interest Statement

The authors have declared no competing interest.

### Clinical Protocols

https://wwwnc.cdc.gov/eid/article/26/8/20-0679_article

